# Identifying barriers and facilitators to physical activity for people with scleroderma: A nominal group technique study

**DOI:** 10.1101/19004432

**Authors:** Sami Harb, Julie Cumin, Danielle B. Rice, Sandra Peláez, Marie Hudson, Susan J. Bartlett, Alexandra Roren, Daniel E. Furst, Tracy M. Frech, Christelle Nguyen, Warren R. Nielson, Brett D. Thombs, Ian Shrier, SPIN-PACE Patient Advisory Team

**Author notes:** Corresponding Author: Ian Shrier, Jewish General Hospital; 3755 Côte-Sainte-Catherine Road; Montreal, Quebec, Canada; H3T 1E2. Sami Harb, Jewish General Hospital; 4333 Côte-Sainte-Catherine Road; Montreal, Quebec, Canada; H3T 1E4. Julie Cumin, Jewish General Hospital; 4333 Côte-Sainte-Catherine Road; Montreal, Quebec, Canada; H3T 1E4. Danielle B. Rice, Jewish General Hospital; 4333 Côte-Sainte-Catherine Road; Montreal, Quebec, Canada; H3T 1E4. Sandra Peláez, Jewish General Hospital; 4333 Côte-Sainte-Catherine Road; Montreal, Quebec, Canada; H3T 1E4. Marie Hudson, Jewish General Hospital; 3755 Côte-Sainte-Catherine Road; Montreal, Quebec, Canada; H3T 1E2. Susan J. Bartlett, McGill University; 687 Avenue Des Pins Ouest; Montreal, Quebec, Canada; H3A 1A1. Alexandra Roren, Hôpital Cochin; 27 Rue Du Faubourg Saint-Jacques; Paris, France; 75014. Daniel E. Furst, University of California, Los Angeles; 10833 Le Conte Avenue; Los Angeles, California, USA; 90095. Tracy M. Frech, University of Utah; 30 North 1900 East; Salt Lake City, Utah, USA; 84108. Christelle Nguyen, Hôpital Cochin; 27 Rue Du Faubourg Saint-Jacques; Paris, France; 75014. Warren R. Nielson, Lawson Health Research Institute; 750 Base Line Road East; London, Ontario, Canada; N6C 2R5. Brett D. Thombs, Jewish General Hospital; 4333 Côte-Sainte-Catherine Road; Montreal, Quebec, Canada; H3T 1E4. Ian Shrier, Jewish General Hospital; 3755 Côte-Sainte-Catherine Road; Montreal, Quebec, Canada; H3T 1E2.

## Abstract

**Purpose:** To identify physical activity barriers and facilitators experienced by people with systemic sclerosis (SSc; scleroderma).

**Materials and Methods:** We conducted nominal group technique sessions with SSc patients who shared barriers to physical activities, barrier-specific facilitators, and general facilitators. Participants rated importance of barriers and likelihood of using facilitators from 0-10, and indicated whether they had tried facilitators. Barriers and facilitators across sessions were subsequently merged to eliminate overlap; edited based on feedback from investigators, patient advisors, and clinicians; and categorized.

**Results:** We conducted nine sessions (n=41 total participants) and initially generated 181 barriers, 457 barrier-specific facilitators, and 20 general facilitators. The number of consolidated barriers (barrier-specific facilitators in parentheses) for each category were: 14 (61) for health and medical; 4 (23) for social and personal; 1 (3) for time, work, and lifestyle; and 1 (4) for environmental. There were 12 consolidated general facilitators. The consolidated items with ≥ 1/3 of participants’ ratings ≥ 8 were: 15 barriers, 69 barrier-specific facilitators, and 9 general facilitators.

**Conclusions:** People with SSc reported many barriers related to health and medical aspects of SSc and several barriers in other categories. They reported facilitators to remain physically active despite the barriers.

**Implications for Rehabilitation:**

- People with scleroderma experience difficulty being physically active due to the diverse and often severe manifestations of the disease, including involvement of the skin, musculoskeletal system, and internal organs.
- In addition to regular care of scleroderma-related symptoms, patients overcome many exercise challenges by selecting physical activities that are comfortable for them, adjusting the intensity and duration of activities, adapting activities, and using adapted equipment or other materials to reduce discomfort.
- Rehabilitation professionals should help people with scleroderma to tailor activity options to their capacity and needs when providing care and advice to promote physical activity.

## Introduction

Regular physical activity is recommended to enhance health among people in the general population [1,2] and for those with chronic diseases [3]. For people with autoimmune rheumatic diseases, health benefits of physical activity training programs may include reduced inflammation, better clinical outcomes, and improved health-related quality of life [4].

Systemic sclerosis (SSc; scleroderma) is a rare chronic, autoimmune rheumatic disease characterized by abnormal fibrotic processes and excessive collagen production that can affect the skin, musculoskeletal system, and internal organs, including the heart, lungs, and gastrointestinal tract [5,6]. People with SSc are classified as having limited (skin involvement of face, neck, and areas distal to the knees and elbows) or diffuse cutaneous SSc (skin involvement proximal and distal to the knees and elbows or trunk). Patients with the diffuse subtype typically have earlier onset of internal organ involvement and more rapidly progressive disease [7].

Most people with SSc can perform aerobic and resistance exercise safely, and regular physical activity is often encouraged [8]. Many, however, face barriers to being physically active [9]. Barriers to being active likely differ across people with SSc, but common barriers may include limitations in physical mobility, respiratory problems, gastrointestinal problems, fatigue, pain, and depression and anxiety [10-12].

Interventions to promote physical activity have been shown to increase activity levels in both the general population [13] and among people with chronic diseases [14]. No physical activity interventions have been developed and tested for people with SSc, and no studies have assessed SSc-specific barriers and facilitators to physical activity. The objective of the present study was to identify barriers and facilitators to physical activity as experienced by people with SSc in order to help rehabilitation therapists provide appropriate care and to generate survey items to be used in a survey that will guide the development of a physical activity promotion program for people with SSc.

## Materials and Methods

### Participants and Procedures

We conducted a series of 90-120 minute face-to-face nominal group technique (NGT) sessions at provincial and national SSc patient conferences in Canada and the United States, and at an international SSc patient conference in France. The NGT was originally designed to structure group discussions so that participants can share and compare experiences and reach consensus [15,16]. More recently, it has been used as a method for directly generating items for needs assessment surveys, including in SSc [17].

For each NGT session, we attempted to recruit up to 8 participants. Eligible participants had received a diagnosis of SSc, were ≥18 years of age, and were fluent in English or French, depending on the conference setting. Prior to each patient conference, we recruited participants through online announcements to participants in the Scleroderma Patient-centered Intervention Network (SPIN) Cohort, a large international SSc cohort; emails and website posts from SSc patient organization partners; and social media (Twitter and Facebook). People with SSc who expressed interest in the study were contacted via email by the study coordinator to confirm eligibility and to provide them with details about the study. At each conference, we also recruited via a table and direct investigator-patient contact. All participants provided written consent and were given the opportunity to ask questions about the study. This study was approved by the Research Ethics Committee of the Jewish General Hospital in Montreal, Quebec, Canada.

Prior to beginning each NGT session, participants completed a brief questionnaire to obtain information on sex, age, race/ethnicity, relationship status, educational attainment, occupational status, SSc diagnosis subtype, and years since SSc diagnosis. Participants were also asked to select the physical activities that they perform from a list (walking, jogging, aerobics, swimming, cycling, yoga or similar exercises) and to add activities if they were not listed. For each activity, they indicated the usual amount of time spent (number of months per year and hours per week).

#### Nominal Group Technique Protocol

We adapted a NGT topic guide from a previously successful NGT study (see Supplemental Material 1) [17]. Before the first NGT session, investigators pilot tested the adapted NGT topic guide. NGT sessions were held in private hotel conference rooms and were moderated by two study investigators who were knowledgeable about SSc and had previous experience with discussion-based research. The moderators for each session always included a doctoral student in clinical psychology (DBR) and either a research assistant (JC), a master’s student in psychiatry (SH), or a clinical psychologist (BDT). The final number of NGT sessions was determined based on the redundancy and consistency of data obtained.

Participants were informed that the objectives of the NGT session were to: (1) develop a list of key barriers to physical activity that they have experienced related to SSc, and (2) develop a list of possible facilitators to overcome the barriers to promote and support physical activity among people with SSc. Participants were first presented with the question: “Think about those barriers or challenges that you have experienced related to SSc. What barriers have you experienced when thinking about or actually being physically active?” They were asked to individually write on a piece of paper, without consultation with other group members, their personal list of examples of barriers to physical activity. Then, they were invited to share one barrier at a time from their lists in a round-robin format until all barriers from each participant’s list had been shared. They were instructed not to repeat barriers that were verbatim to barriers provided by others but to share any barriers that seemed to differ, even if only minimally. If clarification was necessary, moderators used probes to gain a clearer understanding of the barriers shared (e.g., “can you elaborate on that?”). As they were shared, barriers were simultaneously typed on a computer by one moderator and projected onto a screen to be viewed by the moderators and participants. Once all barriers had been shared, moderators led an interactive discussion of the barriers among participants to reword unclear barriers, add any new barriers, remove or merge overlapping barriers, or separate individual barriers with multiple components into more than one barrier. Barriers were revised based on group feedback until agreement was reached for decisions on all barriers.

Next, participants were presented with the second research question: “Think about possible facilitators or strategies to overcome these barriers to promote and support physical activity among people with SSc. What barrier-specific facilitators would be helpful to overcome each barrier, and what general facilitators would be helpful to overcome multiple barriers and address physical activity in general?” For instance, the barrier-specific facilitator example of “electric heated gloves” could address the barrier of “Raynaud’s phenomenon (cold, wind, and humidity)”, whereas the general facilitator example of “exercising with other people” could apply to multiple barriers and physical activity in general. Participants were asked to write any examples of possible barrier-specific and general facilitators, and the same sharing and discussion process used for answering the first research question was then applied to this research question.

Once a final list of unique barrier and facilitator examples was agreed upon, one moderator printed a copy of the list for each participant. In all sessions, participants were asked to independently rate the importance of each barrier on a scale from 0 to 10, with 0 representing barriers that were not personally important to them when thinking about or being physically active, and 10 representing barriers that were extremely important to them when thinking about or being physically active. They rated the likelihood that they would use each barrier-specific facilitator to overcome the barrier to be physically active on a scale from 0 to 10, with 0 representing facilitators that they would not likely use at all, and 10 representing facilitators that they would very likely use. Using the same scale, they also rated the likelihood that they would use each general facilitator. In all but the first two sessions, participants indicated whether they had or had not tried each facilitator.

### Data Processing, Revision, and Analysis

Sociodemographic characteristics and physical activity levels of participants were presented descriptively. All barrier and facilitator examples generated across NGT sessions were compiled into a single list of examples. Many barriers and facilitators identified from individual NGT sessions were similar to those identified in other sessions. Therefore, similar barriers and facilitators were merged into single items, and a merged initial list of items was generated by consensus among investigators. For instance, the barrier item of “difficulty grasping objects” could capture multiple participant examples (e.g., “difficulty gripping weights or bars” and “difficulty grasping things with my hands”).

The initial list of items received three stages of revision by (1) study investigators, (2) 9 members of the SPIN Patient Advisory Team, and (3) 23 health care providers affiliated with SPIN (12 rheumatology physicians, 5 internal medicine physicians, 2 psychologists, 2 physiotherapists, 1 physical and rehabilitation medicine physician, and 1 vascular physician). First, investigators reworded unclear items and excluded items that were too vague (e.g., get a cleaner) or not directly related to physical activity (e.g., surgery), which could not inform the development of a physical activity intervention in SSc. Following this, patient advisors and then health care providers made recommendations to reword items, exclude items, and add new items. Patient advisors and health care providers evaluated whether barrier items met two criteria: (1) they would affect some people with SSc meaningfully (versus only trivially) and (2) they would plausibly be a reason why people with SSc do not participate in physical activity, and whether facilitator items met three criteria: (1) they would be feasibly and realistically used by some people with SSc, (2) they would plausibly address the corresponding barrier (general facilitators would plausibly address multiple barriers and physical activity in general) to support physical activity, and (3) they could be accessed or safely applied by many people with SSc. Study investigators used an iterative process at each stage to implement suggested revisions until consensus on a final list of items was attained.

To group together barriers that share a common basis, we performed a qualitative content analysis [18] of barriers using four categories described by Lascar et al. [19]: (1) health and medical; (2) social and personal; (3) time, work, and lifestyle; and (4) environmental. Study investigators reviewed and attained consensus on classification of barrier items. All processing and analyses were conducted with Microsoft Excel version 16.16.

## Results

Between September 2017 and September 2018, nine NGT sessions were held at the 2017 Scleroderma Society of Nova Scotia Patient Education Forum (one session; Halifax, Canada); 2017 Scleroderma Foundation Tri-State Chapter Fall Research Forum (one session; New York, USA); 2018 Systemic Sclerosis World Congress (two sessions; Bordeaux, France); 2018 Scleroderma Foundation National Patient Education Conference (four sessions; Philadelphia, USA); and 2018 Scleroderma Canada National Conference (one session; Calgary, Canada). The number of participants per session ranged from 3 to 8. Eight sessions were in English and one in French (Bordeaux, France).

### Participant Characteristics and Engagement in Physical Activity

A total of 41 people with SSc (34 female, 7 male) participated in the nine NGT sessions (table 1). Participants ranged in age from 27 to 76 years (mean = 56.2 years; standard deviation = 12.2). Most participants were retired (34.1%), employed full-time (22.0%), or on disability (19.5%). The majority were diagnosed with diffuse SSc (58.5%).

**Table 1.** Participant characteristics.

| **Variable** | **Participants (n=41)** |
| --- | --- |
| **Female, *n* (%)** | 34 (82.9) |
| **Age in years, *mean (standard deviation)*** | 56.2 (12.2) |
| **Race/ethnicity,<sup>a</sup> *n* (%)** |  |
| **White** | 35 (85.4) |
| **Black** | 3 (7.3) |
| **Asian** | 2 (4.9) |
| **Hispanic or Latino** | 2 (4.9) |
| **Aboriginal** | 1 (2.4) |
| **Relationship status, *n* (%)** |  |
| **Never married** | 7 (17.1) |
| **Married or common law** | 29 (70.7) |
| **Separated or divorced** | 5 (12.2) |
| **Highest Level of Education, *n* (%)** |  |
| **Primary school** | 1 (2.4) |
| **Secondary school** | 5 (12.2) |
| **Some college or university** | 12 (29.3) |
| **College or university degree** | 13 (31.7) |
| **Postgraduate degree** | 10 (24.4) |
| **Occupational status,<sup>b</sup> *n* (%)** |  |
| **Unemployed** | 3 (7.3) |
| **Part-time employed** | 3 (7.3) |
| **Full-time employed** | 9 (22.0) |
| **Homemaker** | 1 (2.4) |
| **Retired** | 14 (34.1) |
| **On leave of absence** | 4 (9.8) |
| **On disability** | 8 (19.5) |
| **Systemic sclerosis subtype, *n* (%)** |  |
| **Localized** | 1 (2.4) |
| **Limited** | 13 (31.7) |
| **Diffuse** | 24 (58.5) |
| **Unknown** | 3 (7.3) |
| **Time in years since systemic sclerosis diagnosis, *n* (%)** |  |
| **0 to 5** | 15 (36.6) |
| **5.1 to 10** | 9 (22.0) |
| **> 10** | 17 (41.5) |

<sup>a</sup> Participants could select more than one race/ethnicity. One participant identified as White and Aboriginal, and another identified as White and Hispanic or Latino.
<sup>b</sup> Participants could select more than one occupation. One participant reported being on disability and part-time employment.

All but one participant reported performing at least one type of physical activity (table 2). Most participants engaged in gentle aerobic exercises such as walking, yoga, and swimming. Participants also reported that they performed other physical activities not stated in the questionnaire (n=22) such as tennis, skiing, and gardening.

**Table 2.** Participant engagement in physical activity.

| Type of physical activity | Number of participants (n=41) | Number of months per year<br>*mean (standard deviation),*<br>*range<sup>a</sup>* | Number of hours per week<br>*mean (standard deviation),*<br>*range<sup>a</sup>* |
| --- | --- | --- | --- |
| Walking | 34 | 10.5 (2.6), 4 to 12 | 5.1 (3.6), 0.25 to 14 |
| Yoga or similar exercises | 16 | 11.5 (1.3), 8 to 12 | 4.1 (3.7), 1 to 14 |
| Swimming | 13 | 7.6 (3.9), 3 to 12 | 2.6 (1.5), 0.5 to 5 |
| Cycling | 11 | 9.0 (3.3), 4 to 12 | 3.3 (2.9), 0.75 to 9 |
| Aerobics | 10 | 11.1 (2.0), 6 to 12 | 2.7 (1.2), 1 to 5 |
| Jogging | 5 | 10.4 (3.6), 4 to 12 | 7.9 (6.8), 2 to 16.5 |
| Other physical activity | 22 | 10.1 (3.6), 1 to 12 | 3.9 (2.9), 0.67 to 10 |

<sup>a</sup> If participants stated more than one other physical activity, the data refer to the activity that was performed most frequently per year.

### Barriers and Facilitators to Physical Activity

The nine NGT sessions generated an initial list of 181 examples of physical activity barriers, 457 examples of barrier-specific facilitators, and 20 examples of general facilitators experienced by participants, including similar examples shared in different sessions (see Supplemental Material 2). Figure 1 illustrates the steps used to derive the final survey items from participant examples. Similar examples were merged to attain an initial list of items comprised of 48 barriers, 299 barrier-specific facilitators, and 14 general facilitators. There were 109 facilitator items that participants described as barrier-specific which were deemed as general, merged with existing general facilitator items, and used to inform the description of the 14 initial general facilitators. There were 28 excluded barrier items (most were not directly related to physical activity), 116 excluded barrier-specific facilitator items (most were vague, potentially harmful, or not generally accessible), and 4 excluded general facilitator items (vague or not generally accessible). Additionally, 17 new barrier-specific facilitator items and 2 new general facilitator items were recommended and added. Therefore, the final list of items (see Supplemental Material 3) consisted of 20 barriers, 91 barrier-specific facilitators, and 12 general facilitators. The number of barrier-specific facilitator items per barrier item ranged from 2 to 10.

**Figure 1.**
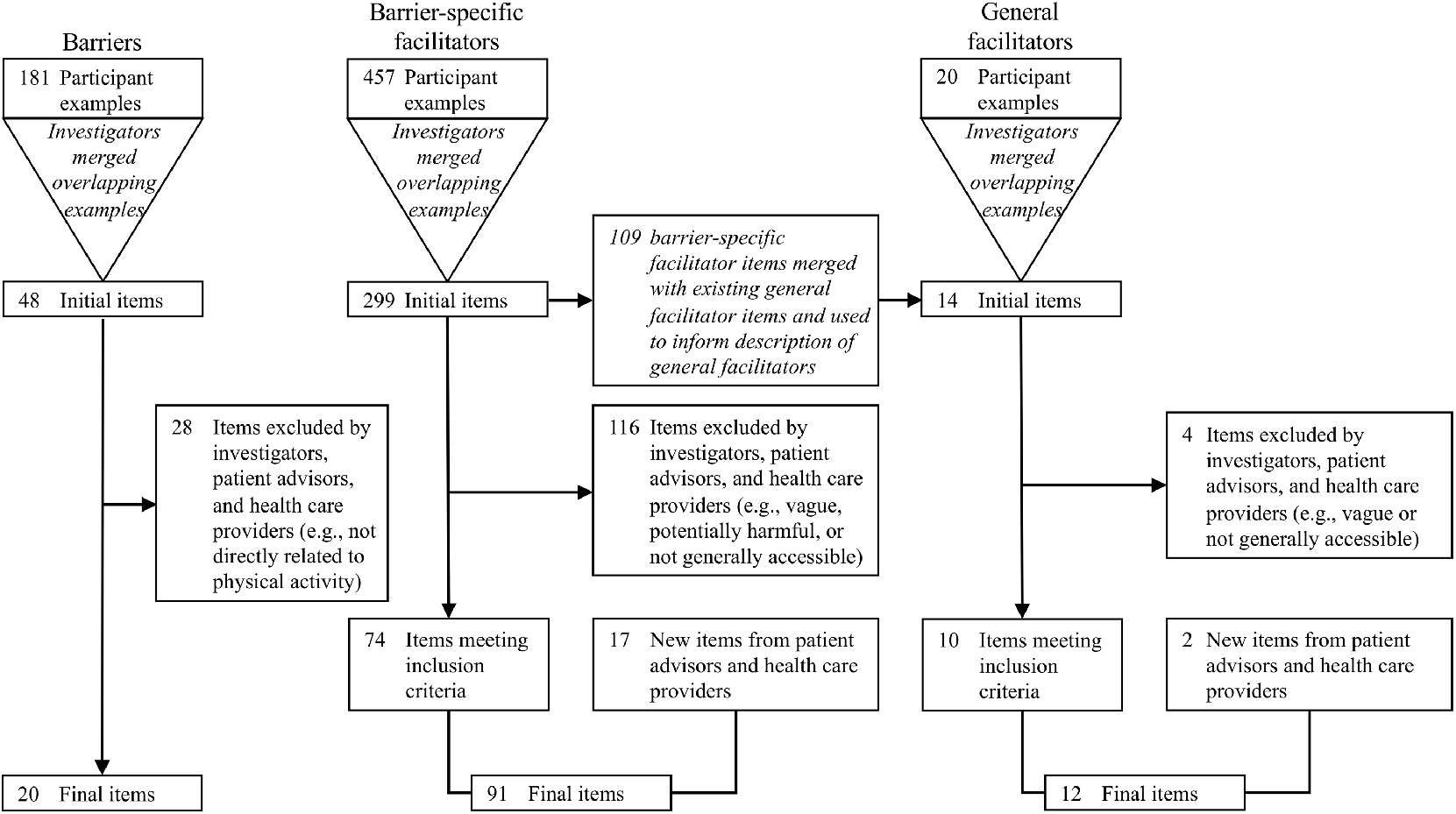
Flow diagram from participant examples to final items.

Of 20 total barriers in the final list of items, 14 (70%) were health and medical barriers which addressed symptoms (e.g., fatigue) as well as medical conditions (e.g., Raynaud’s phenomenon) and activity restrictions (e.g., activities involving water may worsen condition of hands or skin on other areas of the body). Of 61 health and medical barrier-specific facilitators, most involved strategies to beginning and selecting physical activities (e.g., exercise at a time of day when you have the most energy – fatigue barrier), adapting the conduct of activities (e.g., for acid reflux, modify exercise positions to keep your body upright – gastrointestinal problems barrier), adjusting the intensity and duration of activities (e.g., lower the intensity of the exercise to not experience shortness of breath – shortness of breath barrier), using adapted equipment or other materials to reduce discomfort (e.g., insert warmers in gloves or mittens or socks – Raynaud’s phenomenon barrier), and health behaviours to reduce the impact of barriers (e.g., do gentle stretching and movement to warm up the joints before exercise – joint stiffness and contractures barrier).

There were 4 social and personal barriers (20% of total barriers) that addressed feelings about physical activity (e.g., fear of injury or extended recovery time) and being in social settings (e.g., feeling embarrassed or discouraged due to physical ability, appearance, or judgement from others). Of 23 social and personal barrier-specific facilitators, most were methods to feel comfortable with physical activity (e.g., have an introductory session with a qualified exercise trainer to discuss your fears and get an assessment – fear of injury or extended recovery time barrier) and to increase physical activity (e.g., keep an exercise log to track your progress – lack of motivation and difficulty committing to exercise barrier).

There was 1 time, work, and lifestyle barrier related to one’s life circumstances (finding time available to schedule exercise). There were 3 related facilitators, which included exercising at home or work to eliminate travel time, and breaking the exercise into several short periods (also listed as a facilitator to address the fatigue barrier) if one long period was not feasible because of family, work and so on. Lastly, there was 1 environmental barrier related to preventing access to physical activity opportunities (costs related to exercise) with 4 facilitators about free exercise resources and opportunities (e.g., sign up for free activities or exercise classes organized by your community).

Figure 2 shows the distribution of ratings of importance of barrier items from the NGT sessions. The number of ratings per item depended on the number of sessions that identified examples related to the item, the number of examples of experiences related to the item in those sessions (some sessions elicited multiple examples captured by one item), and the number of participants in the sessions where those examples were elicited. The 3 most-rated barriers were health and medical barriers: (1) fatigue, (2) joint stiffness and contractures, and (3) shortness of breath. The 3 most-rated barrier-specific facilitators were also in the health and medical category: (1) wear heated or non-heated warm gloves or mittens and socks (Raynaud’s phenomenon barrier), (2) get enough sleep and plan to take a nap during the day (fatigue barrier), and (3) do strength training exercises (muscle weakness and difficulty with mobility barrier). The 3 most-rated general facilitators related to adapting physical activity were: (1) consult with your health care provider or exercise professional to discuss any concerns and/or custom design an exercise program that is matched to your capacity and needs, (2) exercise at a pace or intensity that is comfortable for you - start easy, progress slowly - if you have pain, adapt the exercise or seek advice, and (3) adapt the exercise or try a new exercise. There were 15 barrier items, 69 barrier-specific facilitator items, and 9 general facilitator items for which at least 1/3 of ratings were ≥ 8 for importance (barriers) or likelihood of using them (facilitators). In addition, there was 1 barrier item, 60 barrier-specific facilitator items, and 9 general facilitator items for which at least 50% of ratings were ≥ 8.

**Figure 2.**
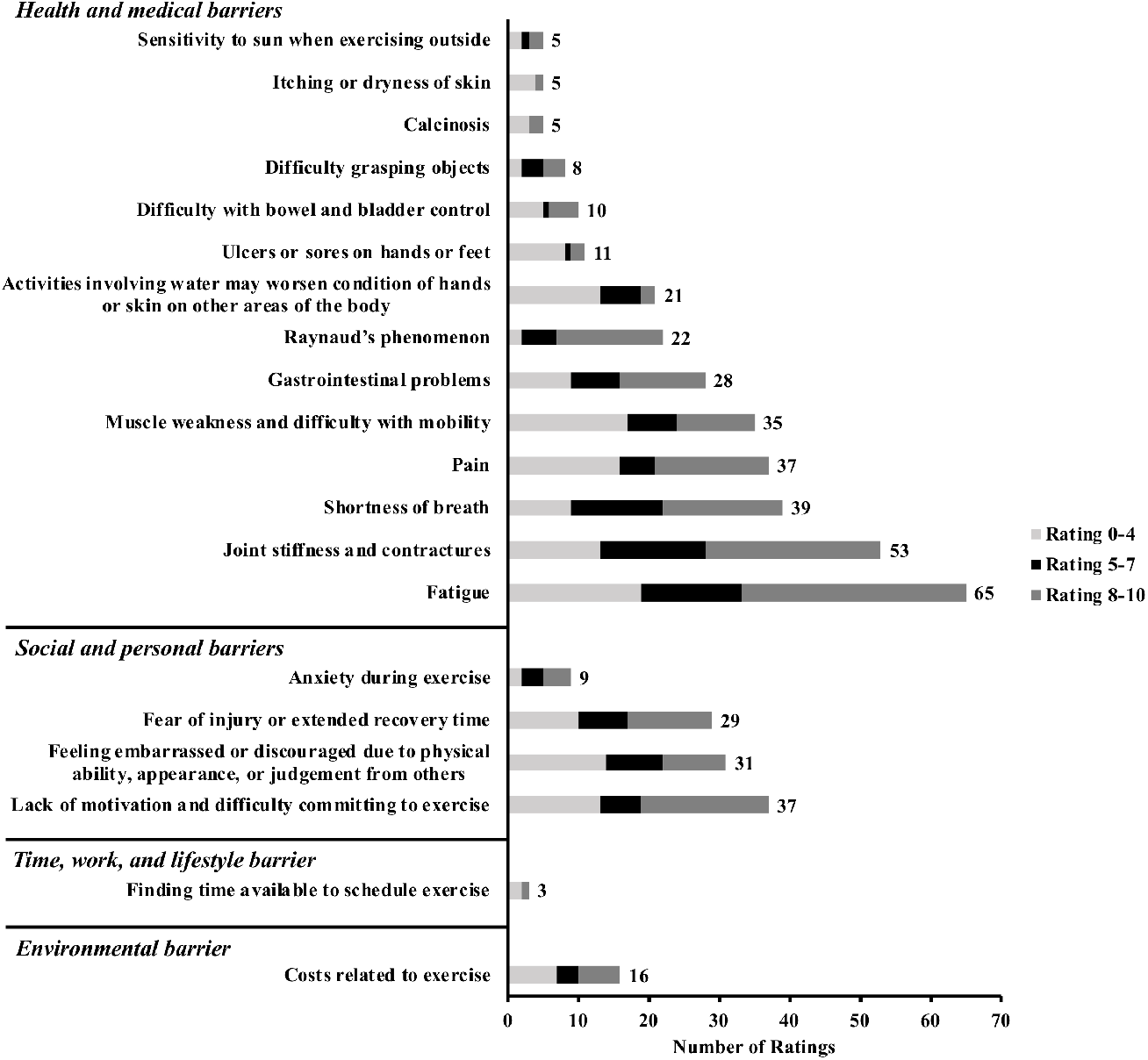
Distribution of ratings for barriers. Numbers to the right of each bar indicate the number of ratings for participant examples captured by the corresponding barrier item. Examples were only rated by participants in sessions where the examples were elicited. The number of ratings can exceed the total of 41 participants because examples were rated before we merged them together into single items. If 3 examples subsequently merged into a single item were each rated by 6 people, then the merged item would have 18 ratings.

## Discussion

Using the NGT method, we identified a list of survey items comprised of 20 barriers, 91 barrier-specific facilitators, and 12 general facilitators to physical activity as experienced by people with SSc. Most barriers fell into the health and medical category, but there were also others grouped into three categories: social and personal; time, work, and lifestyle; and environmental. The list contains previously identified barriers and facilitators from studies of other patient groups or the general population, as well as SSc-specific barriers and facilitators not previously identified in the literature.

Participants’ sociodemographic characteristics were mostly similar to the larger SSc population [20,21]. Specifically, 82.9% were female (versus 87.8% of the SPIN Cohort), mean age was 56.2 (versus 55.5), 85.4% were white (versus 83.6%), 70.7% were married (versus 67.7%), mean years of highest level of educational attainment was 15.7 (versus 15.4) and mean years since diagnosis was 10.8 (versus 9.7). They differed on full- or part-time employment (29.3% of participants versus 40.6% of cohort) and diffuse SSc subtype (58.5% of participants versus 41.3% of cohort).

Barriers in the health and medical category were generally related to SSc disease manifestation or pathology; 12 of 14 health and medical barriers reflect symptoms common in SSc [10]. This study was the first to elicit barriers and facilitators to physical activity directly from people with SSc, but the health and medical barriers identified are consistent with results from other studies that have found that decreased physical activity is associated with fatigue, pain, muscle weakness, and disability in SSc [9,22]. They are also similar to barriers to physical activity reported by people with other autoimmune rheumatic diseases, including fatigue, pain, stiffness, joint symptoms, and reduced mobility or functional ability [23-26].

Barriers in the social and personal; time, work, and lifestyle; and environmental categories were similar to perceived barriers reported by people in the general population and people with other autoimmune rheumatic diseases. These included fear of injury, cost of exercise, lack of motivation, and lack of time [23-29]. One social and personal barrier, feeling embarrassed or discouraged due to physical ability, appearance, or judgement from others, may more closely reflect the experience of people with SSc, including visible changes to their appearance [30].

General facilitators, such as individually adapted physical activity, exercise partners, group exercise, and support from exercise instructors and health care providers, were similar to those identified by people with other autoimmune rheumatic diseases [23-26]. Previous studies have not elicited facilitators to address SSc-specific problems; therefore barrier-specific facilitators identified in the present study will be useful to address these barriers.

Information on barriers and facilitators has been used to develop physical activity interventions for the general population [31] and for people with other diseases (e.g., rheumatoid arthritis [32]). The present study was the first phase of the SPIN-Physical ACtivity Enhancement (SPIN-PACE) Project, the aim of which is to develop, test, and disseminate free-of-charge an online SSc-specific intervention to promote and support physical activity. Based on the results of the present study, a survey will be administered via the SPIN Cohort, an international cohort of over 1,800 people with scleroderma. This will provide information on how common the barriers identified in the present study are and their importance in hindering or impeding engagement in physical activity, as well as the likelihood of using the proposed barrier-specific and general facilitators that were identified. The survey results will inform the development of the planned intervention so that it addresses barriers experienced by people with SSc.

### Limitations

Interpretation of results should consider study limitations. First, we recruited study participants from among people attending patient conferences, and they may not be representative of all people with SSc. In comparison to the SPIN Cohort, a smaller proportion of participants in our study were full- or part-time employed, and a larger proportion of participants had the diffuse SSc subtype. Second, all but one participant reported performing at least one type of physical activity. It is possible that we did not identify important barriers or facilitators for people with SSc who do not engage in physical activity at all. However, the 17 barrier-specific facilitators and 2 general facilitators added by patient advisors and health care providers affiliated with SPIN likely minimized this limitation. Third, although 41 people with SSc participated in the present study and many barriers and facilitators were similar across NGT sessions, it is possible that some potentially important barriers and facilitators were not identified. Therefore, in our planned survey using the SPIN Cohort, we will also ask respondents to suggest new barriers and facilitators.

### Conclusion

In summary, people with SSc reported many barriers related to health and medical aspects of SSc, as well as social and personal; time, work, and lifestyle; and environmental barriers. They further reported facilitators that have helped them remain physically active despite the barriers. The list of barriers and facilitators will be used to survey a much larger number of people with SSc via the SPIN Cohort, which will inform the development of an online physical activity intervention for people with SSc.

## Data Availability

No additional data are available.

## Acknowledgements

We thank the SPIN-PACE Patient Advisory Team for their ongoing contribution to the SPIN-PACE Project, including their review of survey items and suggestions for new items. We also thank health care providers affiliated with SPIN for their review of survey items and suggestions for new items, including: Alexandra Albert, Ilham Benzidia, Camille Daste, María Martín López, Maureen D. Mayes, Susanna M. Proudman, Sébastien Rivière, Anne A. Schouffoer, Virginia D. Steen, and Fredrick M. Wigley.

This research was supported by a Canadian Institutes of Health Research – Strategy for Patient-Oriented Research Grant (PI Shrier) with partner funding from the Scleroderma Society of Ontario. Mr. Harb was supported by a CIHR Canada Graduate Scholarship-Master’s award. Ms. Rice was supported by a Vanier Graduate Scholarship. Dr. Thombs was supported by a Fonds de Recherche du Québec - Santé researcher salary award.

## References

[1] Reiner M, Niermann C, Jekauc D, et al. Long-term health benefits of physical activity–a systematic review of longitudinal studies. BMC Public Health. 2013;13(1):813.

[2] Warburton DE, Charlesworth S, Ivey A, et al. A systematic review of the evidence for Canada’s Physical Activity Guidelines for Adults. Int J Behav Nutr Phys Act. 2010;7(1):39.

[3] Pedersen BK, Saltin B. Exercise as medicine–evidence for prescribing exercise as therapy in 26 different chronic diseases. Scand J Med Sci Sports. 2015;25:1–72.

[4] Perandini LA, de Sá-Pinto AL, Roschel H, et al. Exercise as a therapeutic tool to counteract inflammation and clinical symptoms in autoimmune rheumatic diseases. Autoimmun Rev. 2012;12(2):218–224.

[5] Seibold JR. Scleroderma. In: Harris ED, editor. Kelley’s textbook of rheumatology. 7th ed. Philadelphia: Elsevier; 2005. p. 1279–1308.

[6] Wigley FM, Hummers LK. Clinical features of systemic sclerosis. In: Hochberg MC, editor. Rheumatology. 3rd ed. Philadelphia: Mosby; 2003. p. 1463–1480.

[7] Valentini G. The assessment of the patient with systemic sclerosis. Autoimmun Rev. 2003;2(6):370–376.

[8] de Oliveira NC, Portes LA, Pettersson H, et al. Aerobic and resistance exercise in systemic sclerosis: state of the art. Musculoskeletal Care. 2017;15(4):316–323.

[9] Azar M, Rice DB, Kwakkenbos L, et al. Exercise habits and factors associated with exercise in systemic sclerosis: a Scleroderma Patient-centered Intervention Network (SPIN) cohort study. Disabil Rehabil. 2018;40(17):1997–2003.

[10] Bassel M, Hudson M, Taillefer SS, et al. Frequency and impact of symptoms experienced by patients with systemic sclerosis: results from a Canadian National Survey. Rheumatology. 2010;50(4):762–767.

[11] Kwakkenbos L, Delisle VC, Fox RS, et al. Psychosocial aspects of scleroderma. Rheum Dis Clin North Am. 2015;41(3):519–528.

[12] Thombs BD, Van Lankveld W, Bassel M, et al. Psychological health and well-being in systemic sclerosis: state of the science and consensus research agenda. Arthritis Care Res. 2010;62(8):1181–1189.

[13] Foster C, Hillsdon M, Thorogood M, et al. Interventions for promoting physical activity. Cochrane Database Syst Rev. 2005(1).

[14] Conn VS, Hafdahl AR, Brown SA, et al. Meta-analysis of patient education interventions to increase physical activity among chronically ill adults. Patient Educ Couns. 2008;70(2):157–172.

[15] Harvey N, Holmes CA. Nominal group technique: an effective method for obtaining group consensus. Int J Nurs Pract. 2012;18(2):188–194.

[16] Delbecq AL, Van de Ven AH, Gustafson DH. Group techniques for program planning: a guide to nominal group and Delphi processes. Glenview, IL: Scott, Foresman and Company; 1975.

[17] Rice DB, Cañedo-Ayala M, Turner KA, et al. Use of the nominal group technique to identify stakeholder priorities and inform survey development: an example with informal caregivers of people with scleroderma. BMJ Open. 2018;8(3):e019726.

[18] Mayring P. Qualitative content analysis. Forum Qual Soc Res. 2000;1(2).

[19] Lascar N, Kennedy A, Hancock B, et al. Attitudes and barriers to exercise in adults with type 1 diabetes (T1DM) and how best to address them: a qualitative study. PLoS One. 2014;9(9):e108019.

[20] Fox RS, Kwakkenbos L, Carrier M, et al. Reliability and validity of three versions of the brief fear of negative evaluation scale in patients with systemic sclerosis: a Scleroderma Patient-Centered Intervention Network cohort study. Arthritis Care Res. 2018;70(11):1646–1652.

[21] Dougherty DH, Kwakkenbos L, Carrier M, et al. The Scleroderma Patient-Centered Intervention Network Cohort: baseline clinical features and comparison with other large scleroderma cohorts. Rheumatology. 2018;57(9):1623–1631.

[22] Liem SI, Meessen JM, Wolterbeek R, et al. Physical activity in patients with systemic sclerosis. Rheumatol Int. 2018;38(3):443–453.

[23] Fongen C, Sveaas SH, Dagfinrud H. Barriers and facilitators for being physically active in patients with ankylosing spondylitis: a cross-sectional comparative study. Musculoskeletal Care. 2015;13(2):76–83.

[24] Mancuso CA, Perna M, Sargent AB, et al. Perceptions and measurements of physical activity in patients with systemic lupus erythematosus. Lupus. 2011;20(3):231–242.

[25] Veldhuijzen van Zanten JJ, Rouse PC, Hale ED, et al. Perceived barriers, facilitators and benefits for regular physical activity and exercise in patients with rheumatoid arthritis: a review of the literature. Sports Med. 2015;45(10):1401–1412.

[26] O’Dwyer T, McGowan E, O’Shea F, et al. Physical activity and exercise: perspectives of adults with ankylosing spondylitis. J Phys Act Health. 2016;13(5):504–513.

[27] Booth ML, Bauman A, Owen N, et al. Physical activity preferences, preferred sources of assistance, and perceived barriers to increased activity among physically inactive Australians. Prev Med. 1997;26(1):131–137.

[28] King AC, Castro C, Wilcox S, et al. Personal and environmental factors associated with physical inactivity among different racial–ethnic groups of US middle-aged and older-aged women. Health Psychol. 2000;19(4):354.

[29] Zunft HF, Friebe D, Seppelt B, et al. Perceived benefits and barriers to physical activity in a nationally representative sample in the European Union. Public Health Nutr. 1999;2(1a):153-160.

[30] Jewett LR, Hudson M, Malcarne VL, et al. Sociodemographic and disease correlates of body image distress among patients with systemic sclerosis. PloS One. 2012;7(3):e33281.

[31] Blair SN, Dunn AL, Marcus BH, et al. Active living every day. Human Kinetics; 2011.

[32] Conn VS, Hafdahl AR, Minor MA, et al. Physical activity interventions among adults with arthritis: meta-analysis of outcomes. Semin Arthritis Rheum. 2008;37(5):307–316.

